# On multifactorial drivers for malaria rebound in Brazil: a spatio-temporal analysis

**DOI:** 10.1101/2021.06.24.21259361

**Authors:** Mario J.C. Ayala, Leonardo S Bastos, Daniel A.M. Villela

## Abstract

**Background:** Malaria incidence in Brazil reversed its decreasing trend when cases from recent years, as recent as 2015, exhibited an increase in the Brazilian Amazon basin, the area with highest transmission of *Plasmodium vivax* and *Plasmodium falciparum*. In fact, an increase of more than 20% in the years 2016 and 2017 revealed possible vulnerabilities in the national malaria-control program.

**Methods:** We studied factors that are potentially associated with this reversal, including migration, economical activities, and deforestation, and weakening of investment in control programs. We analyze past incidences of malaria cases due to *Plasmodium vivax* and *Plasmodium falciparum* with a spatio-temporal Bayesian model using more than 5 million individual records of malaria cases from January of 2003 to December of 2018 in the Brazilian Amazon to establish the municipalities with unexpected increases in cases.

**Results:** We observe an increase in imported cases from border countries in Roraima state and found small effects due to deforestation and change of occupations. Also, an overall funding reduction from 2013 to 2016 happened before an increase in malaria cases in five regions in Amazon basin, markedly for *P. vivax* incidence and especially, in Pará and Roraima States.

**Conclusion:** Urban developments, discontinued funding for control programs, migration from border areas, deforestation activities, and different economic activities such as mining and agriculture appear linked to the rebound on malaria incidence. These multifactorial drivers show that malaria control programs require permanent attention towards elimination.

## Background

Control of malaria transmission depends on multifactorial aspects involving vector control, prophylactic treatment, treatment of infected individuals. But these efforts require permanent attention and also control of exogenous conditions. In the case of Brazil, the country reduced its malaria incidence due to both *Plasmodium vivax* and *Plasmodium falciparum* until the year of 2015 but data from recent years exhibited an increase in notified cases [1]. In fact, an increase of 25% between 2016 and 2017 revealed the vulnerability of these efforts [2, 3].

Amazon basin has an area greater than many countries and a huge species diversity due to the Amazon forest and preserved natural resources. These regions have accounted for 99.5% of cases in the last years [4]. But urban developments, deforestation activities, and different economic activities such as mining pose challenges to a large malaria control program.

Previous studies established a set of plausible factors, related to economic activities and investment [5, 6, 7, 8] but none of them evaluated the unexpected increase of cases from 2016 to 2018 in the Amazon region of Brazil. On the other hand, Amazon region presents divergences in malaria incidence between municipalities and States [9], and some of the previous studies only focused on specific regions.

Here, we analyzed incidence trends from 2003 to 2015 defining an expected trend using a statistical model. This model permits us to have predictions of incidences expected in the following years if the trend had stayed. We compare the observed incidence from 2016 to 2018 in all municipalities in Amazon region to the expected values. We also add a Bayesian model to investigate possible relationships of this increase among multiple factors. We identified unexpected incidences due to investment from the Brazilian Ministry of Health, imported cases, deforestation in specific areas. Other aspects were also analyzed, such as occupations of infected individuals.

## Methods

### Epidemiological data

We employed 5.972.715 individual records of malaria cases from January of 2003 to December of 2018 in the States of Amazonas (AM), Acre (AC), Pará (PA), Amapá (AP), Roraima (RR), Rondônia (RO), Mato Grosso (MT), Maranhão (MA) and Tocantins (TO) (see study area in additional file 4). Each record included: notification date, infection data, notification type (active or passive), notification location, occupation, infection location, examination result, periodic control. This data were provided by The Brazilian Information System of Epidemiological Surveillance (SIVEP) where they mainly recorded clinical-symptomatic cases.

The study included only notifications of cases by *P. vivax* and *P. falciparum*. Exclusion criteria were: periodic-control records after treatment; asymptomatic cases (common in active notification); inconsistent information in dates;

Here, we obtained 3.232.766 observations from 3.970.980 (81.41%) to *P. vivax* and 685.317 observations from 885.115 (77.43%) to *P. falciparum*.

### Population data

We also employed annual records of population and Brazilian map per municipalities from the Brazilian Institute of Geography and Statistic (IBGE) webpage in xls and shp file-format.

### Deforestation data

PRODES project of National Institute of Spatial Research provided the annual deforestation in *Km*^2^ per municipality.

### Investment in control programs

We obtained from Brazilian official press the records of funds destined to malaria programs from the Brazilian Ministry of Health in the period from 2003 to 2018. A number of 38 ordinances in the Brazilian official press describe the amount of funds and States or Municipalities as destinations.

### Bayesian models and incidence prediction

We performed a Bayesian model to represent malaria incidence of 808 municipalities in 192 months (from January of 2003 to December of 2018). Spatial variation was labeled as *i* = 1, …, 808 and temporal variation was labeled as *t* = 1, …, 192. *y*_*it*_ is malaria incidence in municipality *i* at month *t*. We modeled *y*_*it*_ as counts in a Poisson distribution with mean *λ*_*it*_ (see equation 1).

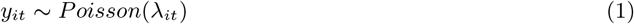

*λ*_*it*_ = *ρ*_*it*_ *ϵ*_*it*_ with *ρ*_*it*_ as incidence rate and *ϵ*_*it*_ as offset; we used municipality population per 100.000 inhabitants as offset. Here, incidence rate is linear predictor in logarithmic scale (equation 2).

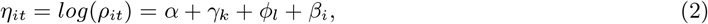

with *α* as average incidence in all municipalities, *γ*_*k*_ as month effect (*k* = 1, …, 12) according with Random Walk Model of order two (RW2) [35], *ϕ*_*l*_ as year effect (*k* = 1, …, 16) according with independent Gaussian random effects (IID) and *β*_*i*_ as municipality effect according with IID. We also implemented a second representation adding an spatial and temporal effect (equation 3).

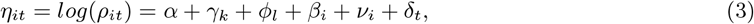

adding *ν*_*i*_ as spatial random effect that consists of two terms according to Besag–York–Mollie (BYM) specification [36], *δ*_*t*_ as temporal random effect derived from the Bernardinelli et al. characterization [37].

We adopted the Integrated Nested Laplace Approximation (INLA) to model estimation [38, 39]. This approach is a Bayesian method that provides computational efficiency through `INLA` package in `R`.

We have two models: a base model (model 1 equation 2) and base model plus spatial and temporal effects (model 2 equation 3). These models were implemented through INLA per each State and then, we evaluated both models using the deviance information criterion (DIC) that assess accuracy of Bayesian models [40]. Model 2 obtained the least DIC for all states (see additional file 5), therefore, we chose this model for predicting using `INLA`.

### Prediction difference

We obtain the difference between real incidence and predicted incidence per month in each municipality. Incidence data from January of 2003 to December of 2015 were employed for modelling and incidence from January of 2016 to December of 2018 were employed for calculating prediction difference. To see impact by year, we calculate an average difference of prediction per year (2016, 2017 and 2018). This produced an estimated difference able to perform a general map by year but it implied an estimation discrepancy (e.g. some municipalities in additional File 3).

Difference maps displayed the average difference of prediction in 2016, 2017 and 2018 for *P. vivax* and *P. falciparum* incidence. We preformed these maps in `R` using lattice package [41].

We defined positive difference as the incidence above prediction, therefore, it represents an unexpected increase of cases above past trends. We selected the most affected municipalities for analysing their positive difference with some variables. Municipalities with a positive difference above 25 cases of *P. vivax* and above 10 cases of *P. falciparum* were selected. Head maps displayed the selected municipalities and they were implemented in `R` using ggplot2 library [42].

### Model with deforestation and occupation categories

Incidence data in individual level also reported the occupation and place of infection (SIVEP data). We counted the frequency of cases from 12 occupations categories per each State in the most affected municipalities with positive difference; occupations categories are agriculture, livestock, housing, tourist, gold-mining, vegetable extraction, hunting/fishing, road building, mining, traveler and other. On the other hand, we counted the frequency of imported cases from border countries per each State in the most affected municipalities with positive difference.

We compared the annual incidence with annual deforestation and the proportion of cases by occupation categories in the period from 2003 to 2018. We calculated the proportion of cases divided by five categories: agricultural activities (agriculture and livestock), outdoor activities (vegetable extraction, hunting/fishing and road building), mining (gold mining and mining), travelling and housing. we performed a Bayesian model to represent malaria incidence of 808 municipalities in 16 years (from 2003 to 2018) adding a random effect of deforestation and each occupation category. Spatial variation was labeled as *i* = 1, …, 808 and temporal variation was labeled as *t* = 1, …, 16. *y*_*it*_ is malaria incidence in municipality *i* at year *t*. We modeled *y*_*it*_ as counts in a Poisson distribution with mean *λ*_*it*_ (see equation 1). Equation 4 joins the structure of equation 3 adding the variable effect before 2016 and after 2015.

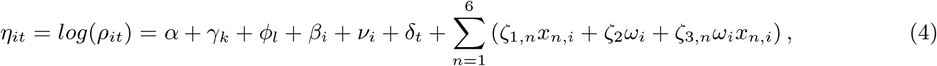

*x*_*n,i*_ represents the variable *n* at year *i, ω*_*i*_ is a binary variable for representing the period from 2016 to 2018 (*ω*_14_, *ω*_15_, *ω*_14_ = 1 and *ω*_*i*_ = 0 for *i <* 14), *ζ*_1,*n*_ is the random effect of variable *n* in all period, *ζ*_2_ is the random effect after 2015 and *ζ*_3,*n*_ is the random effect of variable *n* after 2015. We considered Variables with unexpected increase in their effect on incidence after 2015, variables with *ζ*_3,*n*_ intervals greater than 0.

### Exploratory analysis of investment in control programs

We compared annual malaria incidence with annual investment via descriptive analysis only, since this information is on a timescale (year) different than the one (month) of the statistical model.

## Results

### Model validation and expected incidence after 2015

Analysis from our statistical model permitted to generate samples that describe the time series of malaria incidences due to *P. vivax* and *P. falciparum* across cities and states in the Brazilian Amazon basin. These series can be compared to the actual incidence in all states from 2003 to 2015 for both *P. vivax* and *P. falciparum*. Analysis is carried at municipality levels and presented the aggregate values at state level. Bayesian model estimated increases and decrements in malaria incidence from 2016 to 2018 according to state-past trends and the real incidence were above predictions in some states (Figure 1). This is the case of *P. vivax* incidence in Amazonas (AM), Acre (AC), Pará (PA), Amapá (AP), Roraima (RR), and Rondônia (RO) where real incidence surpassed predictions indicating that these States experimented an unexpected growth of cases. This effect also occurred with *P. falciparum* incidence in AC, AP and RR with less severity than *P. vivax* incidence.

**Figure 1:**
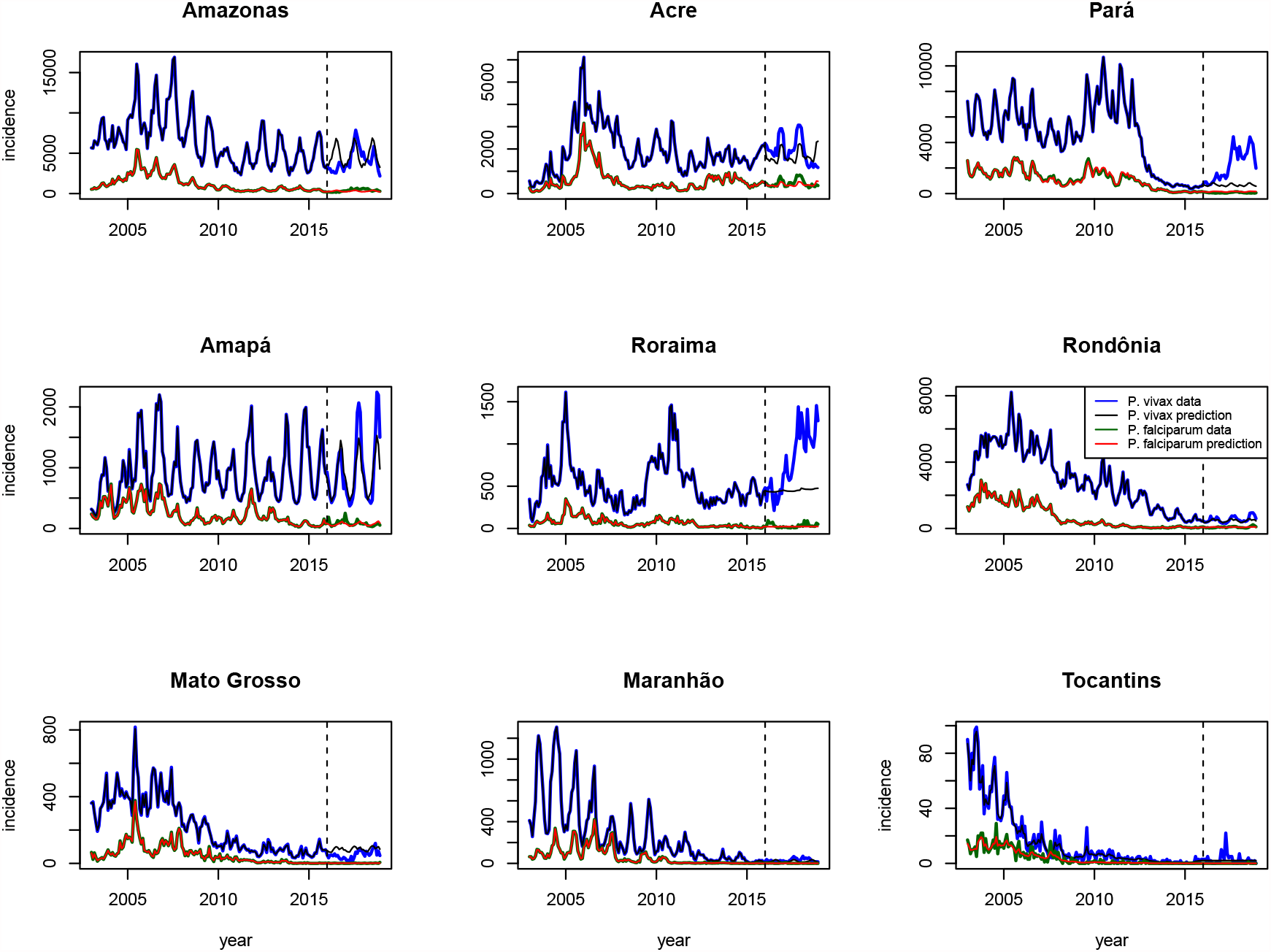
Comparison between model prediction and malaria incidence per state. Blue and black lines represent real incidence and predicted incidence for *P. vivax*; Green and red lines represent real incidence and predicted incidence for *P. falciparum*; Figures illustrate incidence and prediction per month. Dotted line represents the moment (January of 2016) when model started to predict incidence without incidence data.

A set of municipalities presented incidence above predictions -positive difference-where the northwest region of Amazonas (AM), northwest region of Acre (AC), and the northwest region of Rondônia (RO) obtained the highest positive differences for both *P. vivax* and *P. falciparum* incidence (Figure 4). Positive difference for *P. vivax* also involved more regions than *P. falciparum* and five regions contained municipalities with positive difference above 50 monthly cases above prediction: northwest and center region of Amazonas (AM) next to center and south region of Roraima (RR), northeast region of Pará (PA) with border area next to Amapá (AP) (Marajó region), southeast of Pará (PA), northwest region of Acre (AC) and northwest region of Rondônia (RO) (Madeira-Mamore region) next to the south of Amazonas (Purus region).

**Figure 2:**
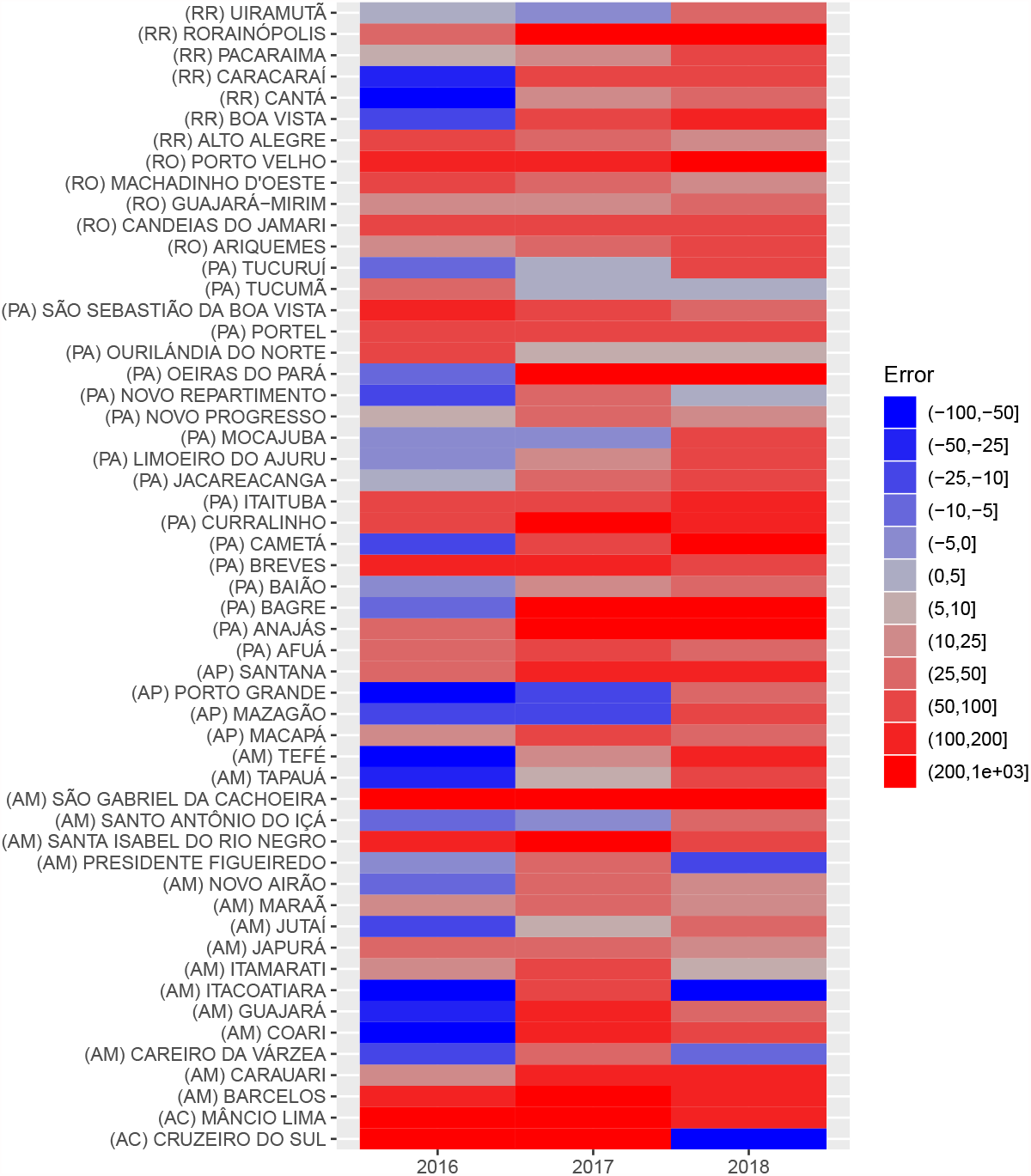
Municipalities with positive difference of prediction (average per each year) above 25 cases for *P. vivax*. The states of Roraima (RR), Rondônia (RO), Pará (PA), Amapá (AP), Amazonas (AM) and Acre (AC) contain these municipalities.

**Figure 3:**
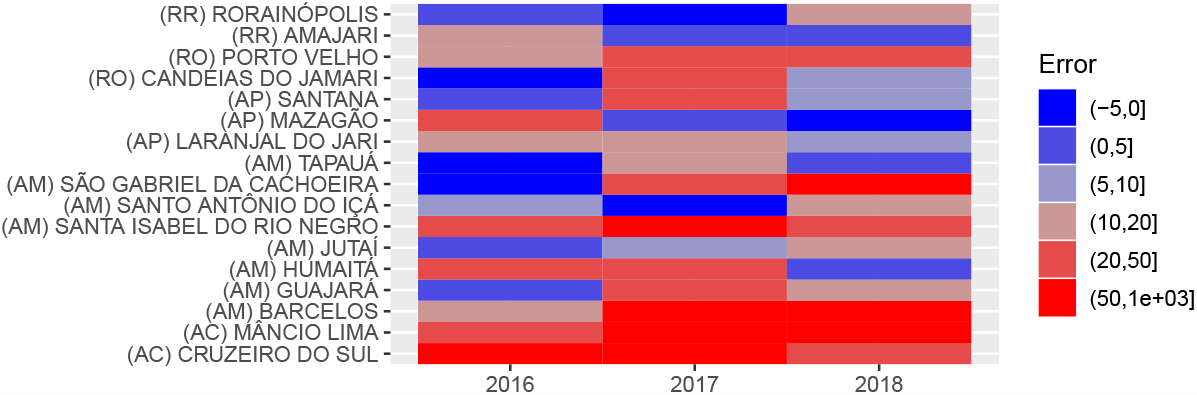
Municipalities with positive monthly difference of prediction (average per each year) above 10 cases of *P. falciparum*. The states of Roraima (RR), Rondônia (RO), Amapá (AP), Amazonas (AM) and Acre (AC) contain these municipalities.

**Figure 4:**
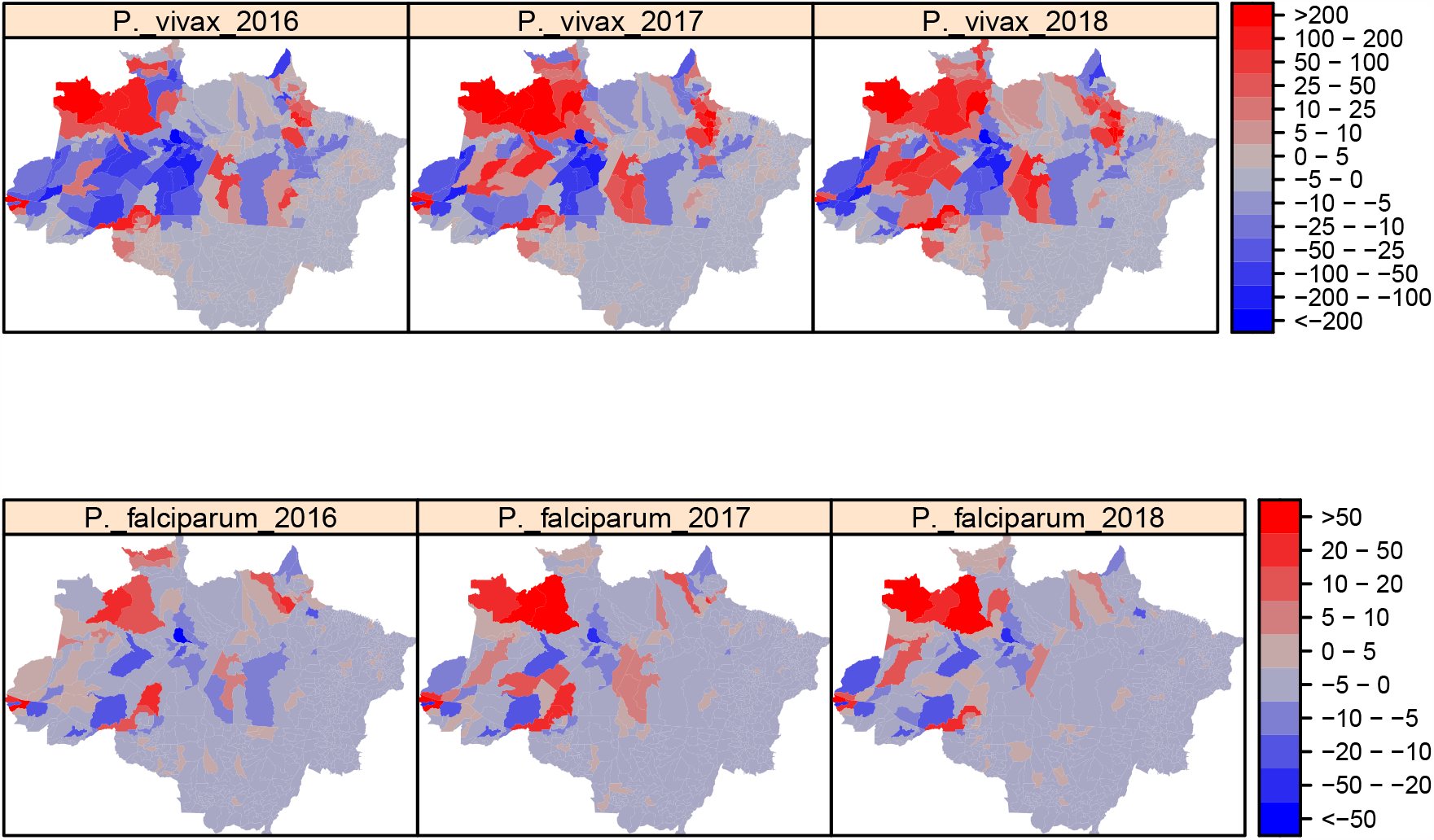
Monthly difference of prediction from 2016 to 2018 (average per each year). Red regions represent municipalities with positive difference (incidence above prediction) and blue regions represent municipalities with negative difference (incidence below prediction). First row displays *P. vivax* prediction difference and the second row displays *P. falciparum* prediction difference.

We found a general rise in the number of municipalities with a positive difference of *P. vivax* from 2016 to 2018 (see Figure 2). The northwest region of AM started with positive differences above 50 in 2016 and then, positive difference extended to all municipalities in the north region, southwest and south-center of AM, and most of municipalities in RR between 2017 and 2018. The northwest region of RO maintained a similar pattern in 4 municipalities and the positive difference in AM extended next to this region in 2018. The positive difference of northeast region in PA and AP extended from 10 municipalities in 2016 (8 in PA and 2 in AP) to 17 municipalities in 2018 (13 in PA and 4 in AP). This also occurred in the southeast of PA where positive difference extended from 2 municipalities in 2016 (2 in PA) to 4 municipalities in 2018 (3 in PA and 1 in AM). AC only maintained the positive difference in two municipalities (Mâncio Lima and Cruzeiro do Sul).

Positive difference in *P. falciparum* exhibited an increase in the northwest region of AM in São Gabriel da Cachoeira, Santa Isabel do Rio Negro and Barcelos (see Figure 3). In general, AM showed an increase in positive difference from 2016 to 2018 whereas AP exhibited a decrease in positive difference. RO and AC maintained positive difference in Porto Velho (RO), Mâncio Lima (AC) and Cruzeiro do Sul (AC).

### Descriptive comparison between variables and prediction difference

Incidence of *P. vivax* and *P. falciparum* exhibited a decrease in all states from 2005 and 2018, except for Roraima exhibiting a peak of *P. vivax* incidence in 2018. This suggests a beneficial effect of investment in malaria control programs. Ministry of Health expanded important contributions from 2011 to 2013 in the most affected states, such as Amazonas, Acre, Pará, Amapá, Roraima and Rondônia that maintained a decrease in malaria incidence on subsequent years. Nevertheless, *P. vivax* incidence went back to an increasing trend in all states starting in 2017, after a diminished investment from October of 2013 to November of 2016. Such rebound effect on *P. vivax* incidence was more dramatic in Pará (PA) and Roraima; and also a rebound effect on the *P. falciparum* incidence present in Amazonas, Acre, Amapá, Roraima and Rondônia, although with less severity.

We also evaluated the impact of imported cases from other countries and occupations in the municipalities that obtained more positive difference of prediction (see Figures 2 and 3). Imported cases only had a relevant impact on *P. vivax* incidence in Roraima (RR) because imported cases were more common from 2017 to 2018 than previous years (see Figure 6); other states did not count an appreciable amount of imported cases. On the other hand, category “other occupations” was the most frequent category (see additional Files 1 and 2). Nevertheless, AM, AC, and PA (only *P. vivax* in PA), reported most of cases as agriculture from categorized occupations for both species. AM and RO also reported an appreciable amount of cases as housing in *P. vivax* cases after 2015. RR had an increase of *P. vivax* cases reported as gold-mining after 2015.

**Figure 5:**
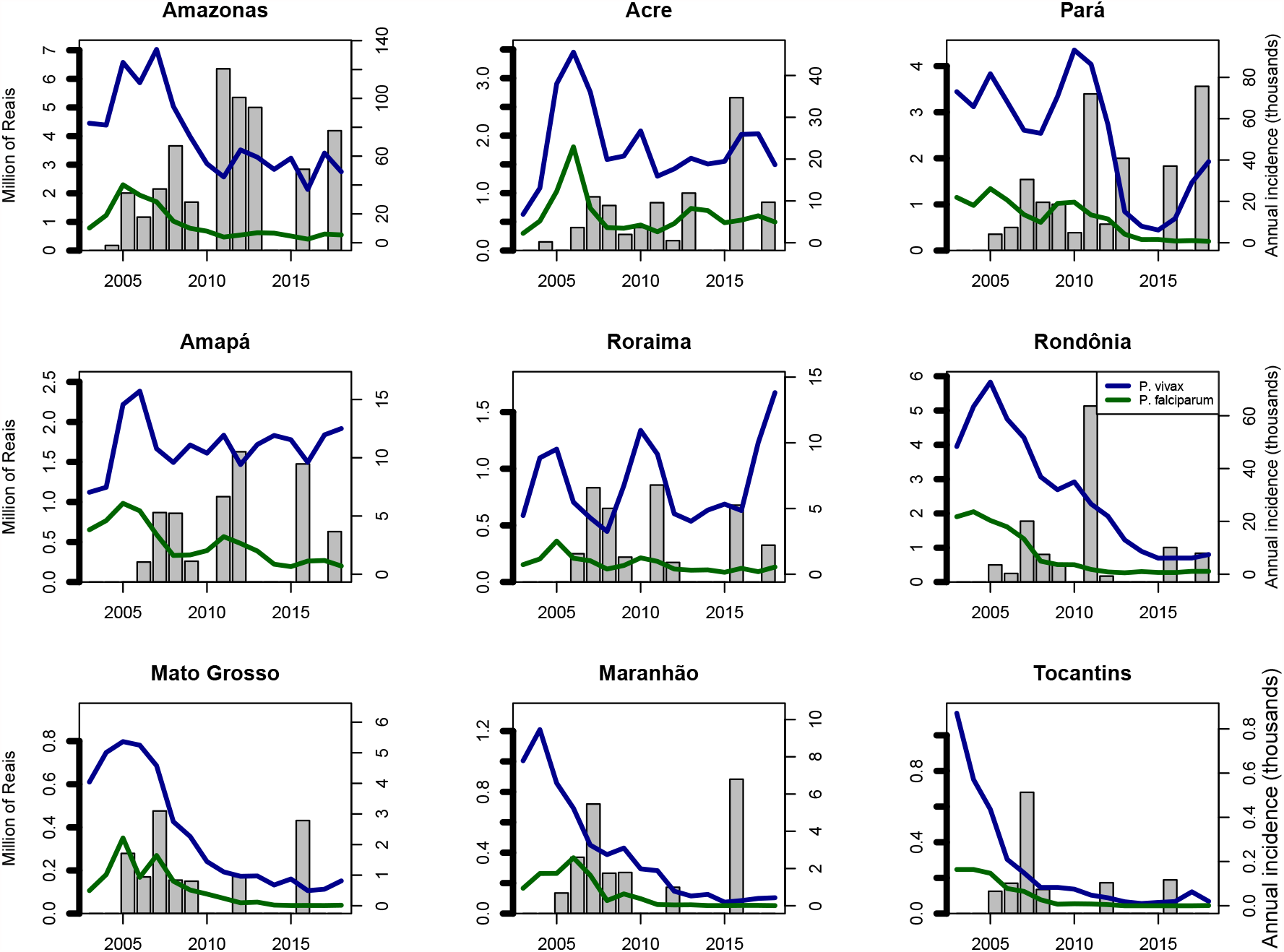
Annual incidence vs annual investment in malaria programmes per state from Ministry of Health. Gray bars represent investment in million of Reais and blue and green lines represent *P. vivax* and *P. falciparum* incidence per year in thousand of cases.

**Figure 6:**
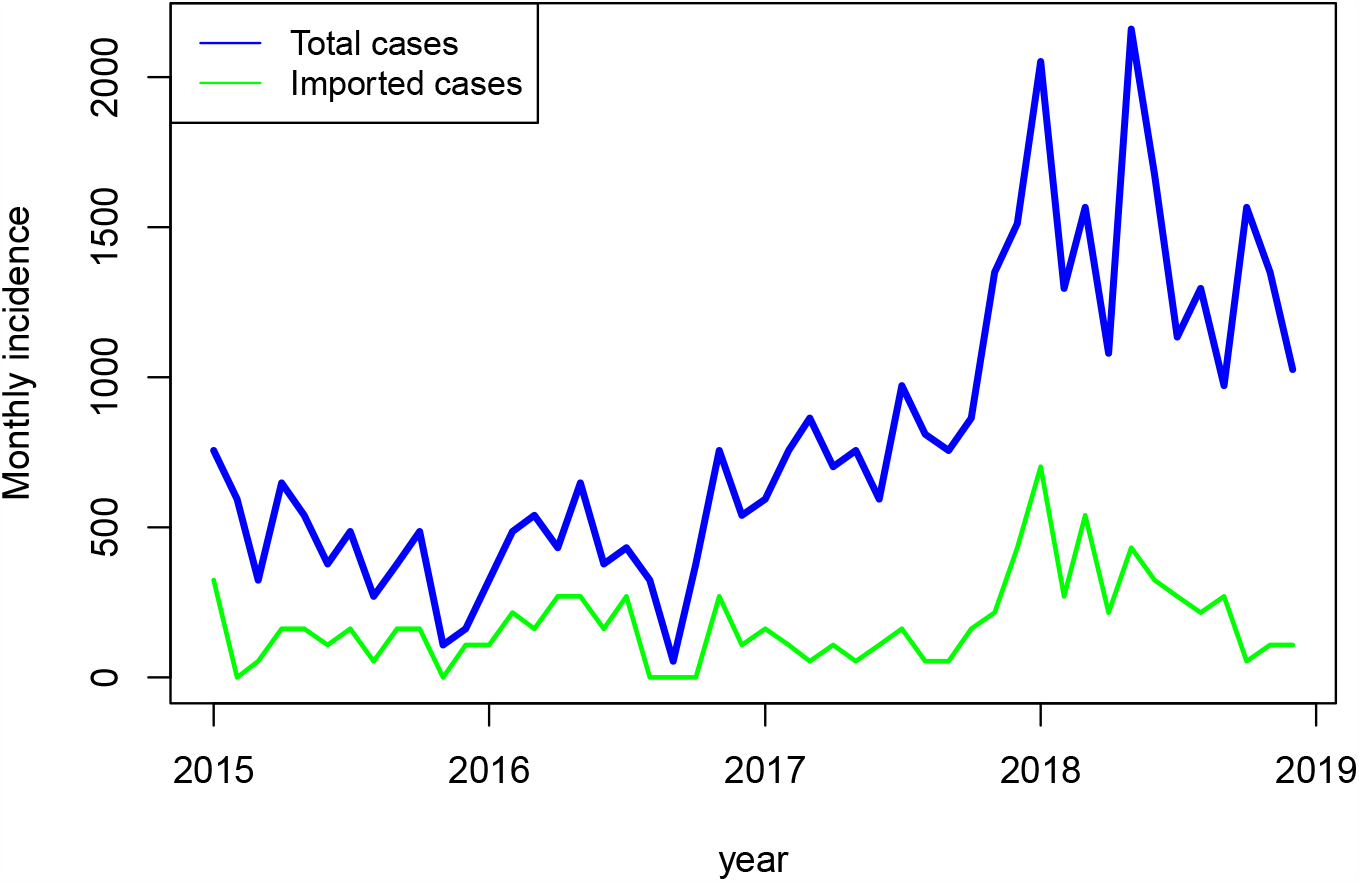
Imported cases in Roraima. Imported cases of *P. vivax* in Uiramutã, Rorainópolis, Pacaraima, Caracarã, Canta, Boa Vista and Alto Alegre in Roraima

We found that the factor of deforestation on incidence only grew up in Roraima (RR) for *P. vivax* after 2015 because of the credibility interval (see table 1). On the other hand, we found a decrease in random effect of deforestation on *P. vivax* incidence in Amazonas (AM) and *P. vivax* incidence in Pará (PA). Thus, deforestation only might explain an unexpected increase in *P. vivax* incidence in Roraima (RR) after 2015 and the effect of deforestation on incidence remained similar in the rest of States after 2015 in comparison to previous years.

**Table 1:** Effects of variables after 2015 (*ζ*_3,*n*_). The mean represents the perceptual increase or decrease on incidence with the increase per variable. Mean in occupations (agricultural, outdoor, mining and traveling) represents the increase in incidence per the increase in 1% of cases reported in each category.

| State | Variable | Species | Mean (95% CI) | Unit |
| --- | --- | --- | --- | --- |
| RR | Deforestation | *P. vivax* | 3.24% (0.30 - 6.26) | $km^2$ |
| RR | Agricultural | *P. falciparum* | 4.53% (2.904 - 6.13) | 1% |
| AM | Deforestation | *P. vivax* | -0.53% (-0.12 - -0.08) | $km^2$ |
| AM | Outdoor | *P. vivax* | 6.21% (0.40 - 12.36) | 1% |
| AM | Agricultural | *P. falciparum* | 3.03% (0.26 - 5.88) | 1% |
| AM | Outdoor | *P. falciparum* | 25.63% (13.15 - 39.51) | 1% |
| RO | Agricultural | *P. falciparum* | 3.21% (0.44 - 6.06) | 1% |
| PA | Deforestation | *P. falciparum* | -0.60% (-1.02 - -0.17) | $km^2$ |
| PA | Agricultural | *P. vivax* | 2.60% (1.32 - 3.90) | 1% |
| PA | Mining | *P. vivax* | 2.36% (0.72 - 4.01) | 1% |
| PA | Outdoor | *P. vivax* | 9.49% (6.33 - 12.75) | 1% |
| PA | Travelling | *P. vivax* | 7.30% (1.92 - 12.95) | 1% |

Table 1 also illustrates the occupations with positive variation in random effect on incidence after 2015 where agricultural, outdoor, mining, and traveling activities might explain the unexpected increase of malaria cases in some states. Agricultural activities might explain the unexpected increase in *P. falciparum* incidence in Rondônia (RO), Roraima (RR), Amazonas (AM), and *P. vivax* incidence in Pará (PA) after 2015. Outdoor activities (hunting/fishing, vegetable extraction, and road building) might explain the unexpected increase in malaria incidence in Amazonas (AM), either *P. vivax* and *P. falciparum* incidence, and *P. vivax* incidence in Pará (PA) after 2015. Mining and traveling activities also might explain the unexpected increase in *P. vivax* incidence in Pará (PA).

## Discussion

The recent trends in malaria incidence from these clinical cases registered in SIVEP-malaria exhibit an increase starting from 2015. The proposed spatio-temporal model has an interaction variable that shows multiple factors interfering with recent trends.

Spatial difference of prediction illustrates regions clusters that concentrated the municipalities with incidence above predictions -unexpected increase in incidence-: northwest and center region of Amazonas (AM) and Roraima (RR), northeast region of Pará (PA) and south of Amapá (AP), southeast of Pará (PA), northwest region of Acre (AC) and northwest region of Rondônia (RO) and south of Amazonas (AM). *P. vivax* incidence above predictions was present in all cluster while *P. falciparum* one was only present in northwest and center region of Amazonas (AM) and Roraima (RR), northwest region of Acre (AC) and northwest region of Rondônia (RO) and south of Amazonas. Carlos et. al found the highest annual parasite index in these regions in the period from 2015 to 2016 and they also found that imported cases between regions and neighbor countries played a role in malaria transmission [24]. Lana et. al also found some clusters in those regions that have provoked the majority of malaria cases in Amazon basin from 2004 to 2018 [43]. Our results revealed the impact of imported cases in Roraima as well Carlos et. al found but we differed in the contribution of imported cases from Perú because we did not find a relation to the unexpected increase on cases in Amazonas and Acre. Imported cases from Venezuelan were the only that impacted malaria transmission in Brazil from neighbor countries. This occurred in the first region in Roraima and the socioeconomic situation in Venezuela might be the principal cause as previous works found [25]. Despite imported cases in Roraima, autochthonous cases represented the principal contributor of cases in Brazil suggesting that local conditions had more impact in the unexpected increase of cases in the majority of clusters.

In fact, socioeconomic situation in Venezuela caused an increase in foreigners seeking for medical attention because the incidence of infectious diseases were higher in Venezuelan population than Brazilian population [26]. Socioeconomic situation in Venezuela also caused an increase in foreigners seeking for job opportunities in gold-mining areas that have presented high risk of malaria transmission [25, 27]. Our results also revealed an increase in cases reported as gold-mining in Roraima from 2015 complementing the difficult situation in Roraima that also accounted an increase in deforestation effect on the incidence of *P. vivax* incidence and an increase in agricultural activities effect on *P. falciparum* incidence.

Brazilian-Amazon basin has a set of environmental, social and economic conditions that might cause unexpected increase in malaria incidence. First, human activities in the Amazon region have induced deforestation that drives in malaria transmission but a recent study also showed a negative feedback where malaria burden reduces forest clearing [28]. Actually, malaria incidence and deforestation has a controversial relation because human development and environmental conditions promote other dynamics increasing or decreasing of malaria burden [29]. Our results showed that deforestation effect had a significant effect on *P. vivax* incidence in Roraima from 2015 to 2018, but not allowing to refute this effect in the unexpected increase of cases in other areas of the Amazon basin.

Second, economic activities as agriculture, mining, tourist, vegetal-products extraction and fishing have boosted malaria in amazon basin. Agrilculture and mining have produced more cases than other economical activities in the central Amazon and the south and west Hiléia settlement (northwest of Brazil) in the 21^*th*^ century [5]. Our results indicated that agriculture were the most reported category in malaria cases in the States of Amazonas and Pará, and agricultural activities also increased their effect in *P. falciparum* incidence in Rondônia, Roraima and Amazonas and their effect in *P. vivax* incidence in Pará. Outdoor activities such as hunting, fishing, vegetable extraction and road building and mining also increased their effect in Pará showing a set of possible drivers of malaria resurgence in this State. The state of Pará has had a constant flow of human migration due to mining, agriculture and infrastructure projects that increase the risk of malaria in this state [30] and this state has shortcomings in health access [31].

We did not find a clear driver of malaria increase in Acre but previous studies found that fish farming in the last decade has provided environmental advantages to *Anopheles* mosquitoes construction in Acre. Priority should be given in health attention [6, 32] and Acre also has experienced an increase in economical activities in periphery zones boosting malaria transmission [8]. In general, our results showed the divergence in possible drivers of malaria rebound because Amazon basin has a epidemiological differences between regions as Canelas et al. found [9].

We found that general funding for the malaria control programs reduced from 2013 to 2016 and this might explain the unexpected increase in malaria cases in Brazil from 2016 to 2018. This occurred to *P. vivax* and *P. falciparum* incidence in all the States where an incidence increase happened. Nevertheless, this effect were more striking in *P. vivax* incidence than *P. falciparum* one, specially, in Pará and Roraima. This revealed that investment withdrawal affected *P. vivax* control more than *P. falciparum* control. *P. vivax* has challenged malaria control programs by difficulties in detection, malaria relapses and earlier transmission [10]. Although *P. falciparum* incidence did not experience similar increases as *P. vivax*, cities in Northwest of Amazonas, Mâncio Lima, Cruzeiro do Sul and Porto Velho presented unexpected increases in *P. falciparum* incidence representing a risk to raise malaria mortality. Weakening in investment had an effect in malaria cases producing a deterioration in health situation as Rondônia exhibited in 1980 [21]. Amazon Basin Malaria Control Project from 1988 to 1986 demonstrated avoiding nearly two million expected-cases and 231.000 expected-deaths in Brazil in spite of low cost-effectiveness of the program [22, 21, 23].

Funding withdrawal for malaria programs poses many limitations to malaria control programs in Latin America, Africa and Asia, even in Brazil. Deterioration in control programs as indoor residual spraying (IRS) entailed a growing incidence in Brazil from 1971 to 1989 [11, 12]. Deterioration in IRS also affected Colombia, Costa Rica, Ecuador, Haiti, Mexico, Panama, Peru and Sri Lanka generating malaria resurgence [7]. Programs in Thailand, India, Kenya, Nigeria, Sudan, Mauritius and Madagascar experimented the resurgence after cutting funds [13, 14, 15, 16, 17, 18]. Our results showed that *P. vivax* incidence increased in 53 municipalities and for *P. falciparum* incidence in 17 municipalities increased over the three years after diminishing investments. For instance, Pará exhibited a clear resurgence after the investment reduction. This failure of maintaining resources to achieve elimination avoiding malaria resurgence implies higher cost in control efforts [19]. Ferreira and Castro advised the necessity of guaranteeing resources for malaria control and elimination to maintain the results to 2015 and avoid resurgence in Brazil [20].

Our analysis with investment funds has a set of limitations. First, the current study only took into the account the resources reported in ordinances from Health Ministry and local health departments in States also provide sources for malaria control that we did not account. Reports from World Health Organization (WHO) suggest other governmental ways to finance malaria in Brazil from 2013 to 2016 [2]. Nevertheless, WHO also notified a reduction in treated cases from 2015 to 2017. Secondly, ordinances defined the expenditure to support public-service of health, epidemiological surveillance, equipment, raw materials, laboratories, emergency care, and general funds but they do not provide information about the specific use of resources and spending periods. This avoids to establish the specific causes in terms of malaria control programs and cost-effectiveness analysis should be fundamental as World Bank did, for the program from 1989 to 1996, as well other countries have done [22, 33]. Third, results compared investment per states without discriminate municipalities of unexpected increases to establish investment priorities; this also happened because Health Ministry has done the majority of resource allocation. Elimination indicators can support investment priorities because only few municipalities concentrate the 80% of incidence and a large number of municipalities in Amazon basin has achieved malaria elimination [34].

## Conclusion

investment reduction from Health Ministry in all States occurred from 2013 to 2016 and this might explain the unexpected increase in malaria cases. This alerts the necessity to maintain sources in malaria control and elimination programs in Brazil. Investment withdrawal can drive in malaria resurgence from the well know risks in the Amazon basin from agricultural activities, migrations, economical activities in periphery zones, mining activities, deforestation and climatic conditions. Maintain resources also provides the support to respond in emergencies like the migratory turmoil in border regions.

## Data Availability

Permission is required to obtain malaria notification data from the Brazilian Ministry of Health.

## Author’s contributions

DV designed and led the project. MC, LB and DV developed the mathematical and statistical models and MC performed the predictions and variable analysis. MC, LB and DV analyzed and discussed the results. MC, LB and DV wrote and approved the final manuscript.

## Availability of data and material

Epidemiological data on malaria cases are available upon request to the Brazilian Ministry of Health.

## Ethics approval and consent to participate

This work involved only access to anonymized data on secondary databases, hence no ethics approval was necessary.

## Consent for publication

Not applicable.

## Acknowledgements

MA and DV are grateful for support from Program Print-Fiocruz-CAPES, Coordenação de Aperfeiçoamento de Pessoal de Nível Superior (CAPES, http://www.capes.gov.br), Conselho Nacional de Desenvolvimento Científico e Tecnológico (Ref. 424141/2018-3) and Fundação Oswaldo Cruz (Fiocruz, http://www.fiocruz.br), and MA is grateful for the scholarship support from Instituto Oswaldo Cruz (IOC, http://www.ioc.fiocruz.br) in the graduate program. DV is a CNPq/Brazil research fellow (Ref. 309569/2019-2).

## References

[1] Pan American Health Organization / World Health Organization. Epidemiological Alert: Increase of malaria in the Americas. 30 January 2018, Washington, D.C.: PAHO/WHO, 2018.

[2] World Health Organization WHO. World Malaria report 2018. France: WHO Library Cataloguing-in-Publication Data, 2018.

[3] Carter KH, Singh P, Mujica OJ, Escalada RP, Ade MP, Castellanos LG, Espinal MA. Malaria in the Americas trends from 1959 to 2011. In Am J Trop Med Hyg, 2015 Feb;92(2):302–316.

[4] Pina-Costa, Anielle de et a. Malaria in Brazil: what happens outside the Amazonian endemic region. Mem. Inst. Oswaldo Cruz [online]. 2014, vol.109, n.5.

[5] Souza PF, Xavier DR, Suarez Mutis MC, da Mota JC, Peiter PC, et al. Spatial spread of malaria and economic frontier expansion in the Brazilian Amazon. In PLOS ONE, 14(6): e0217615, 2019.

[6] Dos Reis, I.C., Codeço, C.T., Degener, C.M. et al. Contribution of fish farming ponds to the production of immature Anopheles spp. in a malaria-endemic Amazonian town. Malar J 14, 452, 2015.

[7] Cohen, J.M., Smith, D.L., Cotter, C. et al. Malaria resurgence: a systematic review and assessment of its causes. Malar J 11, 122, 2012.

[8] Corder RM, Paula GA, Pincelli A, Ferreira MU. Statistical modeling of surveillance data to identify correlates of urban malaria risk: A population-based study in the Amazon Basin. In PLOS ONE, 14(8): e0220980, 2019.

[9] Canelas T, Castillo-Salgado C, Ribeiro H. Analyzing the Local Epidemiological Profile of Malaria Transmission in the Brazilian Amazon Between 2010 and 2015. PLOS Currents Outbreaks, 2018.

[10] Olliaro PL, Barnwell JW, Barry A, et al. Implications of Plasmodium vivax Biology for Control, Elimination, and Research. Am J Trop Med Hyg. 95(6 Suppl):4–14. 2016.

[11] Oliveira-Ferreira J, Lacerda MV, Brasil P, Ladislau JL, Tauil PL and Daniel-Ribeiro Malaria in Brazil: an overview. Malar J, 9:115. 2010.

[12] Marques AC. Human migration and the spread of malaria in Brazil. Parasitol Today, 3:166–170. 1987.

[13] Kondrashin A. World Health Organization.: Epidemiological considerations for planning malaria control in the WHO South-East Asia region. New Delhi: World Health Organization Regional Office for South-East Asia; 1987.

[14] Akhtar R, Learmonth A, Keynes M. The resurgence of malaria in India 1965–76. GeoJournal, 1:69–80. 1977.

[15] Zhou G, Afrane YA, Vardo-Zalik AM, Atieli H, Zhong D, Wamae P, Himeidan YE, Minakawa N, Githeko AK, Yan G Changing patterns of malaria epidemiology between 2002 and 2010 in western Kenya: The fall and rise of malaria. PLoS One, 6:e20318. 2011.

[16] Molineaux L, Gramiccia G. The Garki project: Research on the epidemiology and control of malaria in the Sudan savanna of West Africa. Geneva: World Health Organization; 1980.

[17] Tatarsky A, Aboobakar S, Cohen JM, Gopee N, Bheecarry A, Moonasar D, Phillips AA, Kahn JG, Moonen B, Smith DL, Sabot O. Preventing the reintroduction of malaria in Mauritius: A programmatic and financial assessment. PLoS One, 6:e23832. 2011.

[18] Mouchet J, Laventure S, Blanchy S, Fioramonti R, Rakotonjanabelo A, Rabarison P, Sircoulon J, Roux J. The reconquest of the Madagascar highlands by malaria (in French). Bull Soc Pathol Exot, 90:162–168. 1997.

[19] Rima Shretta, Anton L. V. Avancenã and Arian Hatefi. The economics of malaria control and elimination: a systematic review. Malar J, 15:593. 2016

[20] Ferreira, M.U., Castro, M.C. Challenges for malaria elimination in Brazil. Malar J 15, 284, 2016.

[21] Overholt, Catherine A., Saunders, Margaret K (editors). Policy choices and practical problems in health economics : cases from Latin America and the Caribbean (English). Learning resources series*World Bank Institute (WBI). Washington, D.C. : The World Bank. 1996.

[22] Dariush Akhavan, Philip Musgrove, Alexandre Abrantes, Renato d’A. Gusmão. Cost-effective malaria control in Brazil Cost-effectiveness of a Malaria Control Program in the Amazon Basin of Brazil, 1988-1996 Social Science & Medicine 49; 1385–1399. 1999.

[23] Institute of Economic Research. Cost-benefit analysis for the malaria control program. São Paulo, Brazil. University of São Paulo. 1981.

[24] Carlos BC. Comprehensive analysis of malaria transmission in Brazil. In Pathog Glob Health, Feb;113(1):1–13, 2019.

[25] Judith Recht et al. Malaria in Brazil, Colombia, Perú and Venezuela: current challenges in malaria control and elimination. In Malar J,vol 16, 273, 2017.

[26] Junior MML,Rodrigues GA, Lima MR. Evaluation of emerging infectious disease and the importance of SINAN for epidemiological surveillance of Venezuelans immigrants in Brazil. In Braz J Infect Dis, S1413-8670(19)30418-0, 2019.

[27] PAHO. Plan of action for malaria elimination 2016–2020 (CD55/13). 55th directing council, 68th session of the PAHO regional committte. Washington, D.C.: Pan American Health Organization/World Health Organization; 2016.

[28] Andrew J. MacDonald, Erin A. Mordecai. Amazon deforestation drives malaria transmission, and malaria burden reduces forest clearing. Proceedings of the National Academy of Sciences, 116 (44) 22212–22218. 2019.

[29] Chaves, L.S.M., Conn, J.E., Lopez, R.V.M. et al. Abundance of impacted forest patches less than 5km2 is a key driver of the incidence of malaria in Amazonian Brazil. Sci Rep 8, 7077. 2018.

[30] Lima, Isac da SF, Lapouble Oscar MM, Duarte Elisabeth C. Time trends and changes in the distribution of malaria cases in the Brazilian Amazon Region Mem. Inst. Oswaldo Cruz, 112(1), 8–18. 2017.

[31] Sousa JR, Santo ACF, Almeida WS, Albarado KVP, Magno DL, Rocha JAM, et al. Situação da malária na região do Baixo Amazonas, estado do Pará, Brasil, de 2009 a 2013: um enfoque epidemiológico. Rev Pan-Amaz Saude; 6(4): 39-47. 2015.

[32] Alves Mário Ribeiro, Codeço Claudia Torres, Peiter Paulo Cesar, Souza-Santos Reinaldo. Malaria and fish farming in the Brazilian Amazon Region: a strengths, weaknesses, opportunities, and threats analysis. Rev. Soc. Bras. Med. Trop. 2019

[33] Resign Gunda and Moses John Chimbari. Cost-effectiveness analysis of malaria interventions using disability adjusted life years: a systematic review. Cost Eff Resour Alloc. 15:10. 2017.

[34] Braz, Rui Moreira, and Barcellos, Christovam. (2018). Análise do processo de eliminação da transmissão da malaria na Amazonia brasileira com abordagem espacial da variação da incidência da doença em 2016. Epidemiologia e Serviços de Saúde, 27(3), e2017253. 2018.

[35] Finn Lindgren Håvard Rue On the Second-Order Random Walk Model for Irregular Locations. Scandinavian Journal of Statistics, 35: 691–700, 2008.

[36] Besag J, York J, Mollie A. Bayesian image restoration, with two applications in spatial statistics. Ann Inst Stat Math. 1991;43:1–59.

[37] Bernardinelli L, Clayton D, Pascutto C, Montomoli C, Ghislandi M, Songini. M. Bayesian analysis of space-time variation in disease risk. Stat Med. 1995;14(21–22):2433–43.

[38] Marta Blangiardo, Michela Cameletti, Gianluca Baio, Håvard Rue. Spatial and spatio-temporal models with R-INLA. Spatial and Spatio-temporal Epidemiology. 2013; 7, 39–55.

[39] Rue H. S. Martino, N. Chopin Approximate Bayesian inference for latent gaussian models by using integrated nested laplace approximations J Royal Stat Soc: Ser B (Stat Methodol). 2009; 71 (2), pp. 319–392.

[40] Spiegelhalter DJ, Best NG, Carlin BP, van der Linde A. Bayesian measures of model complexity and fit. Journal of Royal Statistical Society, Series B (Statistical Methodology). 2002; 64:583–639.

[41] Sarkar D. Lattice: Multivariate Data Visualization with R. Springer, New York. 2008, ISBN 978-0-387-75968-5, http://lmdvr.r-forge.r-project.org.

[42] Wickham H. ggplot2: Elegant Graphics for Data Analysis. Springer-Verlag New York. ISBN 978-3-319-24277-4, 2016. https://ggplot2.tidyverse.org.

[43] Lana, R., Nekkab, N., Siqueira, A.M. et al. The top 1%: quantifying the unequal distribution of malaria in Brazil. Malar J 20, 87 (2021). https://doi.org/10.1186/s12936-021-03614-4

