## Supplemental File 1 for "On multifactorial drivers for malaria rebound in Brazil: a spatio-temporal analysis"

### Additional Files

**Additional file 1 — Occupations of *P. vivax*-infected humans in the municipalities with positive average-monthly difference of prediction above 25 cases per month.**

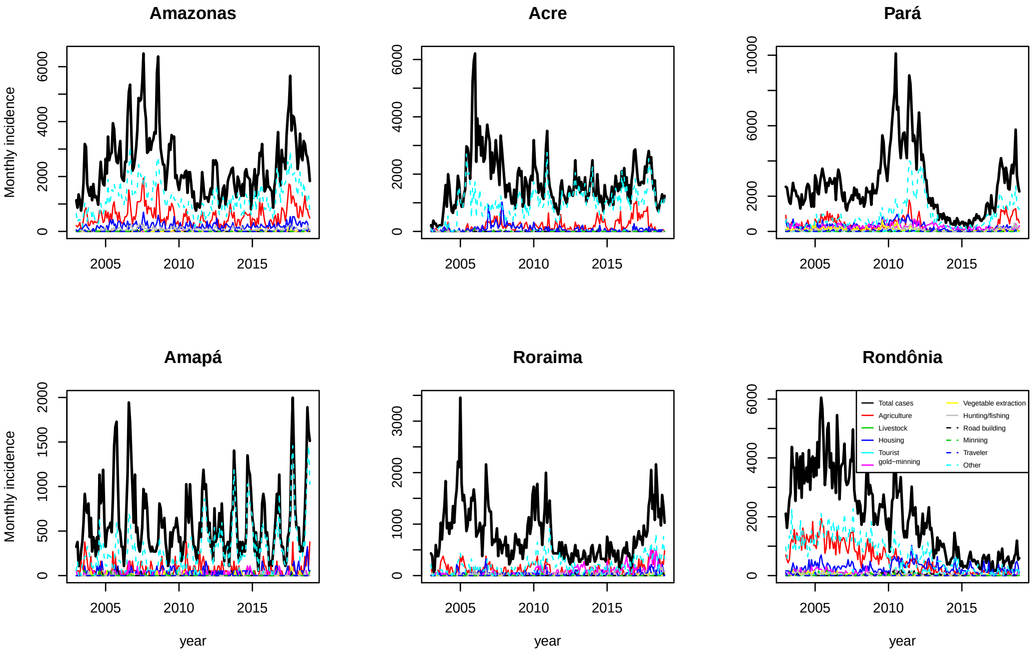


**Figure 1.** Occupations of *P. vivax*-infected humans in the municipalities with positive average-monthly difference of prediction above 25 cases per month.

**Additional file 2 — Occupations of *P. falciparum*-infected humans in the municipalities with positive average-monthly difference of prediction above 10 cases per month.**

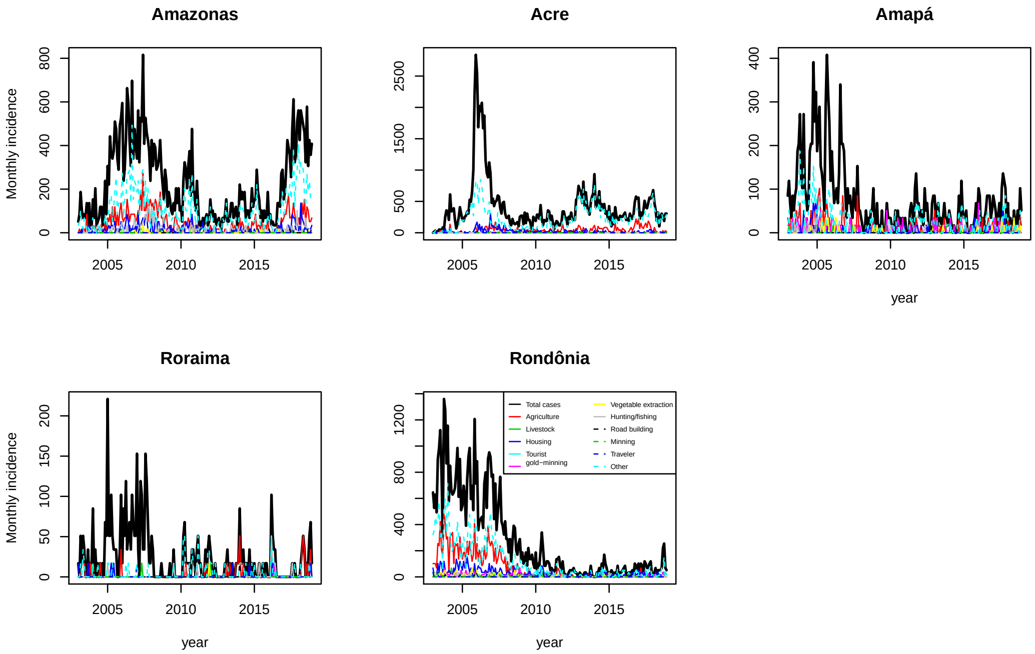


**Figure 2.** Occupations of *P. falciparum*-infected humans in the municipalities with positive average-monthly difference of prediction above 10 cases per month.

**Additional file 3 — Estimation discrepancy in some municipalities. Blue lines represent the prediction difference per month and green lines the average prediction difference per month in each year.**

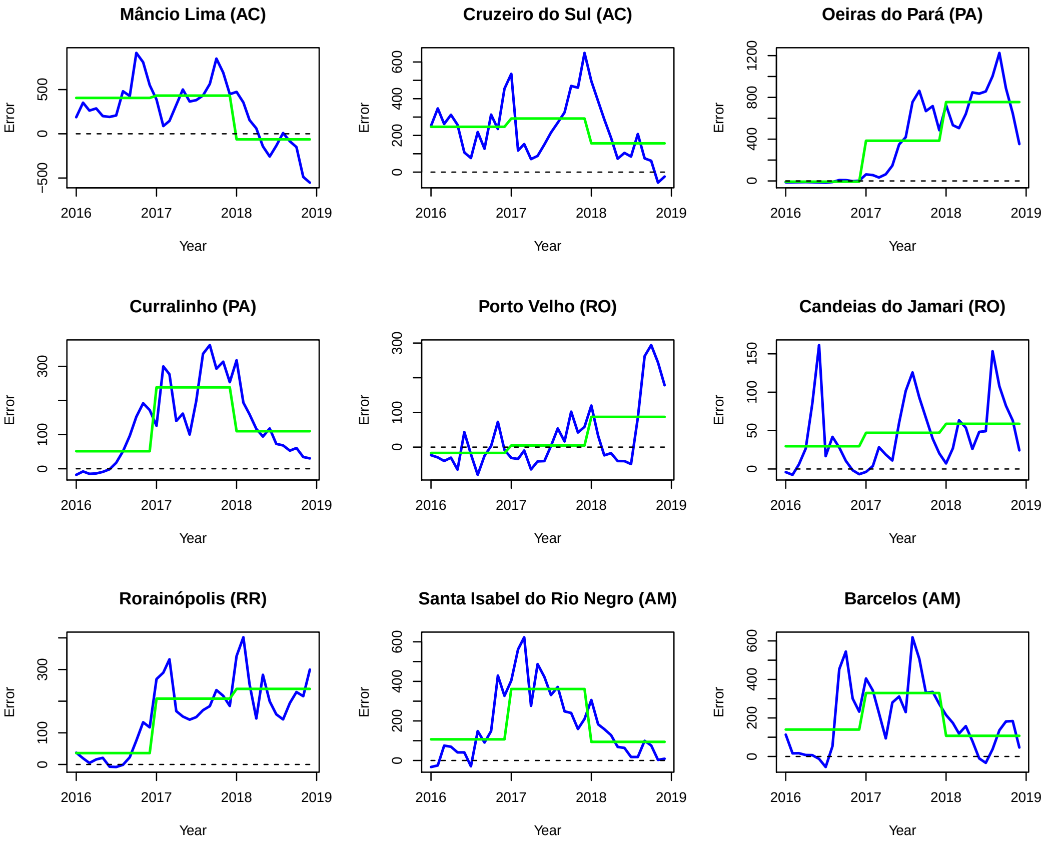


**Figure 3.** Estimation discrepancy in some municipalities. Blue lines represent the prediction difference per month and green lines the average prediction difference per month in each year.

**Additional file 4 — Study area**

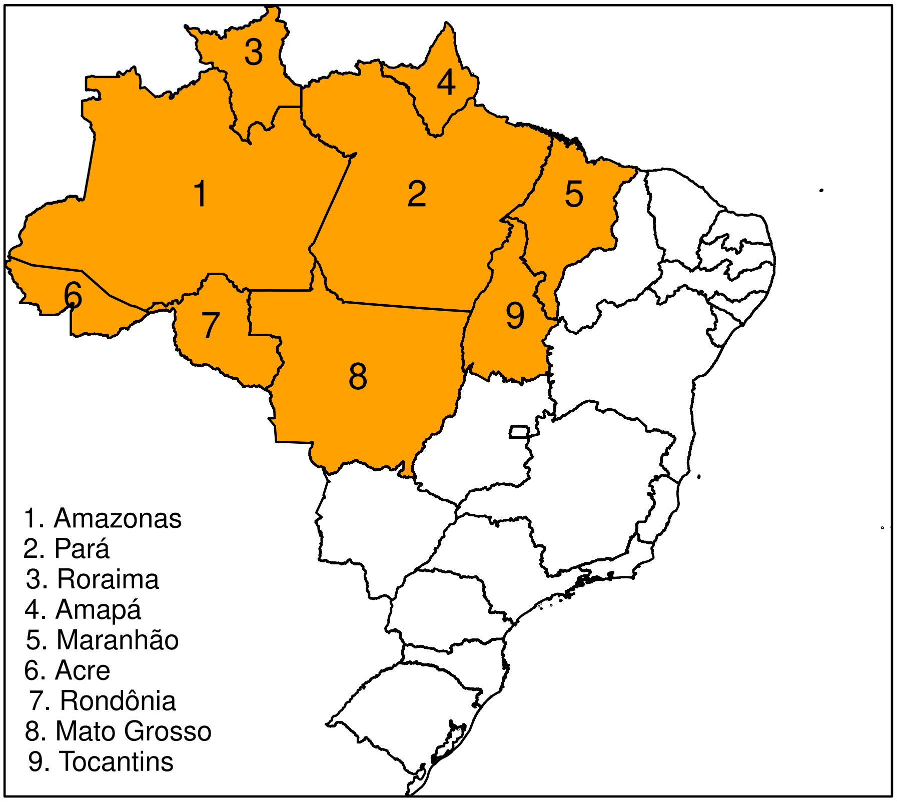


**Figure 4.** Study area: consists in nine states located in Northwest region surrounding by Colombia, Venezuela, Peru, Bolivia, Suriname and French Guiana.

### Additional file 5 — Model choice

**Table 1.** Choice criteria between model 1 and model 2 in all States.

| State | Model | DIC *P. vivax* | DIC *P. falciparum* |
|---|---|---|---|
| AM | Model 1 | 755556.9 | 219725.7 |
| AM | Model 2 | 733638.3 | 215179.9 |
| PA | Model 1 | 691275.9 | 174913.5 |
| PA | Model 2 | 670231.9 | 168723.3 |
| AC | Model 1 | 142410 | 74157.6 |
| AC | Model 2 | 127973.5 | 64414.6 |
| AP | Model 1 | 63918.9 | 34309.1 |
| AP | Model 2 | 59017.5 | 31485.6 |
| RR | Model 1 | 79208.7 | 19119.7 |
| RR | Model 2 | 71863.2 | 18198.3 |
| RO | Model 1 | 140897.5 | 54656.7 |
| RO | Model 2 | 132188.6 | 50360.7 |
| MA | Model 1 | 91185.4 | 31755 |
| MA | Model 2 | 88674.5 | 30904.9 |
| TO | Model | 7342.4 | 2857.4 |
| TO | Model 2 | 7202.2 | 2806.4 |
| MT | Model 1 | 50597.4 | 16439.9 |
| MT | Model 2 | 49091.3 | 15406.9 |
